# Real-World Adaptation and External Validation of Machine-Learning Impedance Spectroscopy for Immediate Postpartum Detection of Obstetric Anal Sphincter Injuries

**DOI:** 10.64898/2026.10.08.26365032

**Authors:** Marcel Młyńczak, Kacper Korzeniewski, Maciej Rosoł, Małgorzata Uchman-Musielak, Ewa Barcz, Magdalena Emilia Grzybowska, Iwona Sudoł-Szopińska, Stephan Kropshofer, Antonino Spinelli, Katarzyna Borycka

## Abstract

Obstetric anal sphincter injury (OASI) requires immediate postpartum recognition for primary repair. Diagnosis relies primarily on clinical examination, as endoanal ultrasound (EAUS) is rarely a bedside option. Therefore, objective diagnostic assessment is most needed during the first postpartum hours. However, it is challenging to obtain the anatomically verified data needed to develop diagnostic technologies for this clinical window. We evaluated the translation of a machine-learning-assisted impedance spectroscopy model into its intended immediate postpartum setting. The model lifecycle comprised three sequential phases: V1 development using anatomically verified data from a prospective multicenter study (N=152); V2 real-world adaptation, comprising post-market data collection, retraining (N=144) to adapt to acute physiological tissue changes, and subsequent separate evaluation (N=125); and prospective external validation of V2 in an unseen cohort (N=70). During this separate evaluation, V2 achieved 89.6% accuracy. In external validation, all examinations occurred within 4 hours of birth (94.3% within the first hour). Against subsequent EAUS, V2 achieved 86.2% sensitivity, 92.7% specificity, and 90.0% overall accuracy. The V2 model was prospectively externally validated in its intended immediate-postpartum setting, supporting ML-assisted impedance spectroscopy as an objective adjunct, and not a replacement, to clinical OASI assessment. Further validation is required to confirm generalizability.

## 1. Introduction

Obstetric anal sphincter injury (OASI) is a clinically important complication of vaginal birth and a major cause of postpartum anal incontinence, with consequences that may persist or become apparent years after delivery [1–3]. Recognition is time-critical: sphincter injury occurs during childbirth and, when identified, primary repair is optimally undertaken in the immediate postpartum period [2]. Diagnosis at this stage relies primarily on systematic clinical examination, including digital rectal examination, but its accuracy depends on examiner experience and clinically relevant injuries, including partial external anal sphincter and internal anal sphincter injuries, may remain unrecognized [2,4,5].

Endoanal ultrasound (EAUS) provides objective anatomical assessment of the anal sphincter complex and is widely used as a reference imaging method for OASI [2,6,7]. Although EAUS can be performed immediately after birth [7], it requires dedicated equipment and expertise and is rarely used as an acute bedside investigation. Consequently, the clinical window in which OASI must be managed does not readily coincide with the timeframe in which objective anatomical reference data can be routinely obtained.

This temporal mismatch also creates an important challenge for the development of machine-learning-based diagnostic technologies. Supervised model development requires reliable reference labels, yet large anatomically verified datasets obtained during the first hours after birth are inherently difficult to generate. Models developed using high-quality postpartum reference data must therefore ultimately be translated to an intended-use environment that may be underrepresented in the original training data [8]. The immediate postpartum period also represents a distinct biological environment, in which acute tissue changes may alter the impedance conditions encountered by the model.

Electrical impedance spectroscopy (IS) offers a rapid bedside approach to assessing tissue properties without requiring anatomical image interpretation. Combined with automated machine-learning (ML) analysis, IS enables classification of impedance patterns associated with sphincter integrity [9,10]. Previous pilot and prospective multicenter studies demonstrated the feasibility and diagnostic performance of ML-assisted IS against EAUS across the postpartum period [9,10]. The prospective multicenter study provided anatomically verified data for initial model development but included women examined across a broader postpartum window extending to eight weeks after birth [9]. Following deployment of the initial model, real-world use provided substantially greater representation of measurements obtained within the immediate postpartum period and identified performance variation concentrated within this early domain. These observations indicated that the intended-use environment was not fully represented by the original development data.

These findings provided the basis for a typical sequential ML model lifecycle [11,12] comprising initial clinical development (V1), adaptation using early-postpartum real-world data (retraining to V2), and prospective external validation of the locked V2 model. The objective of this study was to evaluate the translation of an ML-assisted impedance spectroscopy model from clinical development to its intended immediate-postpartum setting through real-world adaptation and prospective external validation against an anatomical reference standard. In this context, the technology is positioned strictly as an objective adjunct to bedside clinical examination, rather than its replacement.

## 2. Results

### 2.1. Phase 1: Pivotal Clinical Study and Initial Model Performance

The clinically evaluable population of the study comprised 152 women (mean age 31.3 ± 4.6 years; mean BMI 25.0 ± 3.6 kg/m^2^). Based on anatomical integrity confirmed by EAUS, participants comprised 30 women without perineal trauma or OASI (Group A), 61 with first- or second-degree perineal trauma without OASI (Group B), and 61 with third- or fourth-degree tears involving the anal sphincter complex (Group C). Detailed participant characteristics have been reported previously [9].

Each participant underwent two IS measurements. During the prespecified *in silico* modeling phase, 19 measurement runs showing inter-measurement impedance differences >1 kΩ at 1 kHz were excluded. The resulting model-development dataset comprised 279 reproducible impedance measurements: 117 from women with EAUS-confirmed OASI and 162 from women without OASI.

Using repeated 10-fold cross-validation with 10 repetitions and participant-level data partitioning, the resulting artificial neural network (V1 model) achieved a mean accuracy of 90.0% (±0.4%), sensitivity of 90.0% (±1.2%), and specificity of 90.0% (±0.7%) against the EAUS reference standard.

### 2.2. Phase 2: Real-World Model Adaptation and Evaluation

Following deployment of the V1 model into routine clinical use, PMS accumulated real-world measurements from the intended immediate-postpartum setting. Among cases where subsequent diagnostic information was available, 15 discordant outputs were identified, comprising 6 false-positive and 9 false-negative results. These discrepancies occurred predominantly within the first 24h after delivery and were frequently associated with acute postpartum conditions, including marked tissue oedema and recent primary repair sutures. Together, these observations confirmed that the immediate-postpartum measurement environment was underrepresented in the original development dataset and provided the basis for targeted model adaptation.

Retraining and model adaptation resulted in V2, which was finalized and locked before evaluation in the separate two-center dataset of 125 recordings. The confusion matrix for this validation dataset yielded 84 true negatives, 28 true positives, 4 false negatives, and 9 false positives, translating to an overall accuracy of 89.6%, a sensitivity of 87.5%, and a specificity of 90.3%.

### 2.3. Phase 3: Prospective External Validation of the Locked V2 Model

Of the 151 women prospectively enrolled, 70 completed the anatomical reference assessment and formed the external validation cohort. The remaining 81 participants did not complete EAUS follow-up and were therefore not included in the paired validation analysis. Median maternal age was 32.5 years (IQR 29.0–35.0): 42 women (60.0%) were primiparous and 28 (40.0%) multiparous. Instrumental vaginal delivery occurred in 11 cases (15.7%), episiotomy in 27 (38.6%), and neonatal birthweight ≥4000 g in 14 (20.0%).

Impedance spectroscopy was performed within the intended immediate-postpartum window in all 70 participants. Sixty-six examinations (94.3%) were completed within the first hour after delivery, and all were completed within 4 h. Forty-nine examinations (70.0%) were performed by midwives.

EAUS was performed at a median of 28 days postpartum (IQR 19.25–31.75; range 1–40 days). Seven participants (10.0%) underwent EAUS on the first postpartum day, whereas the remaining examinations were performed during subsequent follow-up. EAUS identified OASI in 29 participants (41.4%) and no sphincter injury in 41 (58.6%).

Against EAUS as the anatomical reference standard, V2 correctly classified 63 of 70 participants, corresponding to an overall accuracy of 90.0% (95% CI 80.8–95.1%). Of the 29 EAUS-confirmed OASI cases, 25 were correctly identified and 38 of 41 participants without OASI were correctly classified, yielding a sensitivity of 86.2% (95% CI 69.4–94.5%) and specificity of 92.7% (95% CI 80.6–97.5%). All performance metrics are summarized in Table 1. The clinical diagnostic pathway, including comparison with bedside clinical assessment and analysis of clinical–IS discordance, is reported separately [13].

**Table 1.** Diagnostic performance of the impedance spectroscopy V2 model in the prospective external validation cohort. EAUS served as the anatomical reference standard. Confusion matrix values are presented as numbers of subjects, while performance metrics are reported as percentages or ratios, with confidence intervals in brackets.

| Metric | Impedance spectroscopy V2 model |
| --- | --- |
| True Positive | 25 |
| False Negative | 4 |
| True Negative | 38 |
| False Positive | 3 |
| Sensitivity (95% CI) | 86.2% (69.4–94.5%) |
| Specificity (95% CI) | 92.7% (80.6–97.5%) |
| Accuracy (95% CI) | 90.0% (80.8–95.1%) |
| Positive Predictive Value (95% CI) | 89.3% (72.8–96.3%) |
| Negative Predictive Value (95% CI) | 90.5% (77.9–96.2%) |
| Positive Likelihood Ratio (95% CI) | 11.78 (3.92–35.34) |
| Negative Likelihood Ratio (95% CI) | 0.15 (0.06–0.37) |

Exploratory descriptive performance estimates across selected delivery characteristics revealed consistent diagnostic capability across major clinical variables. The overall accuracy was comparable between women with episiotomies (92.6%) and those without (88.4%), as well as between instrumental (100.0%) and non-instrumental (88.1%) deliveries. Time to examination appeared not to degrade performance, with tests conducted within the first hour yielding an 89.4% accuracy compared to 100.0% accuracy for the few tests (N=4) performed between 1 and 4 hours postpartum. Because of the small numbers within individual subgroups, particularly for instrumental delivery, macrosomia and examinations performed more than 1 h after birth, these analyses were considered descriptive only and were not used to infer subgroup-specific differences in model performance.

## 3. Discussion

This study describes the translation of a machine-learning-assisted impedance spectroscopy model from anatomically verified clinical development to prospective external validation in its intended immediate-postpartum setting. Three findings are particularly important. First, the initial model (V1) achieved approximately 90% diagnostic performance when developed and evaluated using anatomically verified data from the prospective, multicenter study. Second, subsequent routine clinical use provided substantially greater representation of the immediate postpartum measurement environment and enabled targeted model adaptation using real-world data. Third, after this adaptation had been completed and V2 locked, diagnostic performance was maintained in a previously unseen prospective cohort, with accuracy of 90.0%, sensitivity of 86.2%, and specificity of 92.7%, against EAUS. Importantly, all impedance measurements were obtained within 4 h of delivery and 94.3% within the first postpartum hour. Seventy percent of examinations were performed by midwives, indicating that external-validation performance was achieved within a midwife-inclusive maternity workflow. This should not be interpreted as evidence of operator independence, which was not formally assessed, but supports further evaluation across different professional operators and routine postpartum pathways. The close correspondence between initial model-development performance and prospective external-validation performance indicates that diagnostic performance was maintained during translation to the intended-use environment.

This development pathway is particularly relevant to the interpretation of ML-assisted diagnostic technologies. For clinicians, confidence in such a test depends not only on its reported sensitivity and specificity, but also on the clinical relevance of the data used to develop the model and on whether final performance has been demonstrated in patients who played no role in model development [14]. An ML model learns classification patterns from the data represented during training; performance established during development therefore cannot by itself establish equivalent performance in a different population or measurement environment [8]. In the present analysis, the prospective multicenter investigation provided high-quality anatomical labels through EAUS but included women examined across a postpartum interval extending to 8 weeks. Routine clinical use subsequently provided greater representation of the first hours after delivery. Model adaptation through retraining using these data was followed by locking and prospective evaluation in a participant-independent cohort. This separation between model learning and subsequent evaluation is central to the interpretation of the Phase 3 results. The transition from the V1 to V2 should therefore be viewed in the context of translation to the intended clinical domain. Real-world discrepancies occurred predominantly during the first 24h after delivery, with acute postpartum conditions, including marked tissue edema, biological fluids and recent suturing, recurring among affected measurements. These observations are compatible with a change in the impedance measurement environment during the immediate postpartum period and indicated that this domain was insufficiently represented in the original development data. The present study does not establish which individual physiological factors contributed to particular classification errors, nor does it demonstrate that the V2 model specifically distinguishes sphincter disruption from edema or suturing. It does, however, illustrate why representation of the intended-use environment matters for a data-driven diagnostic model. Importantly, after adaptation to this domain, the V2 model maintained approximately 90% overall accuracy when prospectively tested under the same early-postpartum conditions.

This issue is closely linked to a specific challenge in OASI diagnosis. Sphincter injury occurs during childbirth and needs to be recognized during the immediate postpartum period, when decisions regarding primary repair are made. EAUS provides objective anatomical assessment of the sphincter complex but requires dedicated equipment and expertise and is not routinely available as a bedside reference investigation during this same clinical window [2,7]. Consequently, the time at which diagnostic information is most clinically useful does not readily coincide with the time at which objective anatomical reference data can routinely be obtained. This creates an additional challenge for ML development, because reliable anatomical labels are required for supervised model development and validation, whereas large anatomically verified datasets obtained within the first postpartum hours are difficult to generate. The development pathway evaluated here was designed to address this temporal mismatch: anatomically verified data supported initial model development while real-world data increased representation of the immediate-postpartum measurement environment in which the technology is intended to be used.

The clinical relevance of these findings relies on positioning ML-assisted impedance spectroscopy as an objective adjunct to clinical examination, rather than a replacement for bedside assessment or anatomical imaging. Both serve functions that extend beyond the binary classification provided by the model. The potential value of impedance spectroscopy is complementary: it can provide an additional objective diagnostic layer during the immediate postpartum period, when clinical decisions need to be made but anatomical imaging is generally not immediately accessible.

Detailed analysis of the relationship between clinical assessment, impedance spectroscopy, and EAUS, including the added diagnostic information provided by impedance spectroscopy, is reported separately [13]. In the present study, the key finding is that ML-based interpretation maintained its diagnostic performance after translation to the immediate-postpartum environment in which this additional information is intended to be used.

The study also has important limitations. The real-world data used during Phase 2 were obtained during routine clinical use and were not generated within a single prospectively standardized research protocol; their provenance, available reference information, and separation from subsequent evaluation datasets are therefore important when interpreting this phase. The prospective external-validation cohort was relatively small and derived from a single center. Moreover, 81 of 151 enrolled participants did not complete EAUS follow-up, introducing the possibility of selection or verification bias. Recruitment enriched the analyzed cohort for women at increased risk of OASI, resulting in an OASI prevalence of 41.4%; consequently, caution is required regarding the generalizability of these findings, as predictive values cannot be directly extrapolated to an unselected maternity population with lower disease prevalence. EAUS was performed at a median of 28 days postpartum, creating a temporal interval during which postpartum anatomy and, in women undergoing repair, sphincter appearance may have changed. In addition, EAUS was performed by a single highly experienced examiner who was not blinded to the earlier clinical and ONIRY findings. Neurogenic defects were not included [15]. Finally, although measurements were obtained by different professional operators, formal inter-operator reproducibility was not assessed.

These limitations define the next stage of evaluation. The principal priority is broader prospective validation of the V2 model across independent centers, operators and consecutive maternity populations, particularly in lower-prevalence settings representative of routine postpartum screening. Further studies should quantify intra- and inter-operator reproducibility and determine how PASS/REFER information influences subsequent diagnostic pathways, use of confirmatory imaging and, ultimately, patient outcomes.

In conclusion, this study demonstrates the translation of an ML-assisted impedance spectroscopy model from anatomically verified clinical development to prospective external validation in its intended immediate-postpartum environment. Real-world data enabled adaptation to an underrepresented early-postpartum measurement domain, after which the locked V2 model maintained approximately 90% diagnostic accuracy in a previously unseen prospective cohort examined predominantly within the first hour after birth. These findings support ML-assisted impedance spectroscopy as an objective adjunct to immediate postpartum OASI assessment and illustrate a broader principle relevant to ML-assisted diagnostics: clinical confidence depends not only on model performance, but also on representative development data, transparent model adaptation, and independent validation after model deployment.

## 4. Materials and Methods

### 4.1. Study Design and Overview

The paper describes the clinical development, real-world adaptation, and independent external validation of the ML-assisted IS system (ONIRY, OASIS Diagnostics, Warsaw, Poland) intended for the immediate postpartum detection of OASI.

The methodological framework comprised three sequential phases: (1) the initial V1 model development and validation using data from a prospective multicenter clinical study with EAUS as the anatomical reference standard; (2) real-world model adaptation, including retraining using early-postpartum data collected through post-market surveillance (PMS); and (3) prospective external validation of the V2 model in a previously unseen immediate-postpartum cohort against an EAUS reference standard.

All described studies were conducted in accordance with the Declaration of Helsinki.

Phase 1 additionally followed Good Clinical Practice for clinical investigations of medical devices (ISO 14155:2020) and was approved by the ethics committees competent for each study site: Ethics Committee of the Institute for Maternal and Child Care, Prague (1/19.03.2021), the Ethics Committee for Research with Medicines of the Health Areas of León and Bierzo (2186), the Ethics Committee of the University Hospital Brno (47/21Zdrav.), the Bioethics Committee at the District Medical Chamber in Warsaw (KB/1362/21), and the Ethics Committee of AGEL Hospital Košice-Šaca (ONIRY 3/2/2020).

Phase 2 used anonymized data generated during routine clinical use of the CE-marked device and collected through post-market surveillance (PMS). No additional study-specific procedures were performed. OASIS Diagnostics had no access to patient identities or linkage keys and no means of identifying or re-identifying the patients from the supplied data. No separate ethics committee review was sought for these Phase 2 activities. These data were subsequently used for model adaptation in accordance with applicable data-protection and PMS requirements.

The Phase 3 prospective observational study was approved by the Bioethics Committee at the District Medical Chamber in Warsaw (RNN/312/25/KE). As a non-interventional observational study, Phase 3 was not registered in a clinical trial registry.

Written informed consent was obtained from all participants in Phase 1 and Phase 3.

The study and manuscript were reported with reference to STARD-AI 2025 guidelines for reporting diagnostic accuracy studies using artificial intelligence [16] and TRIPOD+AI guidelines for reporting clinical prediction models that use regression or machine learning methods [17]. Completed checklists are provided in the Supplementary Material S1-S2.

### 4.2. ONIRY system and impedance measurement

The ONIRY system (Class IIa medical device under EU MDR 2017/745) comprises an impedance spectrometer, a single-use endoanal probe, and an ML-based interpretation module. The 12-mm-diameter probe contains eight electrodes to acquire impedance measurements around the anal canal without manual rotation. A low-amplitude sinusoidal alternating current (<1 mA) is applied across a frequency range of 1–100 kHz, generating measurements of impedance magnitude, resistance, reactance and phase shift.

A single examination takes approximately one minute. The acquired impedance signals undergo automated quality control to identify conditions incompatible with reliable measurement, including improper probe placement or inadequate tissue contact. Measurements failing predefined quality-control criteria are rejected and the operator is prompted to repeat the examination. Following the Phase 2 model adaptation, the quality-control procedure was additionally updated with a low-impedance alert to identify measurements potentially affected by conditions associated with very low tissue impedance, including marked edema or the presence of biological fluids.

Signal-derived features are parameterized and then processed by the ML interpretation module, which provides an immediate binary output: PASS, indicating no impedance pattern suggestive of OASI, or REFER, indicating an impedance pattern suggestive of OASI and the need for further clinical evaluation. For REFER results meeting the predefined localization threshold, the system additionally displays the estimated location of the abnormal impedance pattern within the anal canal.

The impedance acquisition principle and hardware platform remained unchanged throughout the model development pathway. The principal technological modification concerned the ML interpretation and associated quality-control procedures, with the initial V1 model trained and validated in Phase 1 subsequently adapted using real-world immediate-postpartum data to produce the V2 model trained in Phase 2 and evaluated in Phases 2 and 3.

### 4.3. Phase 1: Pivotal Clinical Study and Initial Model Development

The prospective, comparative, multicenter clinical study involved 152 women examined up to 8 weeks following vaginal delivery (ClinicalTrials.gov identifier: NCT04903977; submitted: 17^th^ May 2021, study start: 20^th^ May 2021, first posted online: 27^th^ May 2021; the registry record was submitted before enrolment of the first participant; its first posting followed the registry’s quality-control review; current status: completed) [9]. The study comprised a clinical data-acquisition phase followed, as prespecified in the protocol, by an in silico modeling phase in which the definitive ML interpretation V1 model was developed using impedance measurements paired with three-dimensional EAUS assessment as the anatomical reference standard.

Raw impedance measurements underwent preprocessing and feature extraction as previously described [9]. Extracted features included statistical characteristics of the impedance signals within predefined frequency ranges. Measurements that did not meet predefined reproducibility criteria were excluded before model development.

An artificial neural network (ANN) was developed and evaluated using a 10-time 10-fold cross-validation procedure. Because multiple measurements could originate from the same participant, all data partitioning was performed at participant level to prevent measurements from the same woman appearing in both training and validation folds. After validation a final ML model was trained using the validated architecture and hyperparameters. The resulting V1 model was locked before its subsequent deployment in clinical use.

### 4.4. Phase 2: Real-World Model Adaptation

To systematically address the dataset shift observed in the delivery-room setting, Phase 2 was structured into three sequential sub-phases: data collection via post-market surveillance (PMS), model retraining, and subsequent evaluation. Given the routine clinical nature of the post-market data utilized in this phase, endoanal ultrasound (EAUS) was not universally available; therefore, the reference standard for these specific real-world datasets was established based on the clinical totality of evidence (incorporating initial bedside assessment, documented anatomical findings, and subsequent clinical management).

#### Phase 2A: Post-Market Surveillance and data collection

Following deployment of the V1 model into routine clinical use, PMS provided an increasing body of real-world measurements from the intended immediate-postpartum setting. Evaluation of these data showed that discordant outputs were concentrated predominantly within the first 24 h after delivery, a period that had been underrepresented in the original development dataset. This real-world evidence provided the basis for targeted adaptation of the model to the intended-use environment.

#### Phase 2B: Algorithm retraining and V2 model finalization

Model retraining utilized a dataset of 144 early-postpartum impedance recordings obtained during routine clinical use of ONIRY at seven clinical centers in Poland, Spain and Austria. Signal processing and feature engineering were revised to provide an updated input representation comprising lesser number of signal-derived features based on impedance magnitude, resistance, reactance, phase shift and local variance. Multiple candidate model architectures were evaluated, with hyperparameter tuning performed exclusively within the development data. The final model was trained and evaluated using a strict 90/10 hold-out split (90% training, 10% separate internal validation). No participant contributed measurements to more than one partition. Model architecture, hyperparameters and the decision threshold were determined without access to the subsequent independent evaluation dataset.

The resulting V2 model comprised an ensemble of two multilayer perceptron (MLP) neural networks whose binary predictions were combined using a logical AND decision rule. The ensemble rule and decision threshold were selected during model development to improve specificity while maintaining sensitivity. The decision threshold was selected to optimize balanced accuracy, while maintaining the prespecified target sensitivity of >80%. Once the architecture, model parameters and decision threshold had been finalized, the V2 model was locked. Specific details regarding the exact MLP architecture, number of layers, and optimized hyperparameters are maintained as proprietary commercial confidential information.

#### Phase 2C: Validation of the Locked V2 Model

The V2 model was subsequently evaluated on a separate dataset comprising 125 recordings obtained from two clinical centers in Poland. This dataset was not used for model training, hyperparameter tuning, architecture selection or threshold determination.

### 4.5. Phase 3: Prospective External Validation of the Locked V2 Model

To evaluate the performance of the V2 model in its intended immediate-postpartum setting, a prospective external validation against an independent anatomical reference standard was performed within an observational cohort study conducted at the Holy Family Specialist Hospital (Warsaw, Poland). The clinical pathway and added diagnostic value of impedance spectroscopy within this cohort are reported separately [13]; for the present analysis, the cohort constituted a previously unseen external validation dataset.

#### Study population

Eligible women were prospectively and consecutively recruited (when clinical circumstances permitted recruitment) following vaginal delivery between October 2025 and May 2026. Women aged ≥ 18 years were eligible if they had at least one predefined OASI risk factor (primiparity, operative vaginal birth, neonatal birthweight ≥ 4000 g, or gestational age ≥ 41 weeks) or if the initial clinical examination raised diagnostic uncertainty regarding possible sphincter injury. Exclusion criteria were inability to provide informed consent, clinical instability following delivery, or a contraindication to rectal examination as determined by the treating clinician. A total of 151 women were enrolled. Of these, 81 did not complete the planned EAUS follow-up and therefore did not contribute to the paired diagnostic accuracy analysis. The external validation cohort comprised the remaining 70 women with both immediate-postpartum impedance spectroscopy and subsequent EAUS data.

#### Clinical workflow

IS examinations were performed as a part of immediate postpartum assessment and completed within 4h of delivery: Of the 70 examinations, 66 (94.3%) were performed within the first postpartum hour, and 49 (70%) were performed by midwives. Standard clinical examination, including digital rectal examination (DRE), was performed by an attending obstetrician or senior trainee. The IS result was available to the treating clinician at the bedside as additional diagnostic information. Clinical management remained at the discretion of the treating team and was not determined by the IS result alone. Participants were not allocated to alternative interventions or comparison groups, and the CE-marked device was used within its intended purpose.

#### Reference standard

All 70 participants underwent a three-dimensional EAUS in a dedicated perineal clinic at a median of 28 days after delivery (cases labelled OASI-positive included both unrepaired and repaired injuries, based on the features visible on EAUS). Examinations were performed by a single expert ultrasonographer with >25 years of experience. Anal sphincter integrity was classified according to the Sultan criteria and EAUS served as the anatomical reference standard [18]. The ultrasonographer was not blinded to the earlier clinical and IS findings.

The Phase 3 cohort was completely independent of all preceding model-development stages. No Phase 3 data were available during Phase 1 or Phase 2 model training, feature selection, hyperparameter tuning, architecture selection or threshold determination. V2 was finalized and locked before prospective Phase 3 evaluation.

### 4.6. Statistical Analysis

Categorical variables are expressed as counts and percentages. Diagnostic performance of V2 was assessed against EAUS as the anatomical reference standard. Sensitivity, specificity, PPV, NPV, overall accuracy, were calculated. Given that PPV and NPV are highly dependent on disease prevalence, Positive Likelihood Ratio (LR+) and Negative Likelihood Ratio (LR−) were additionally calculated as prevalence-independent metrics. Confidence intervals (CI) for sensitivity, specificity, PPV, NPV and accuracy were calculated using the Wilson score intervals. CI for LR+ and LR− were calculated using the logarithmic method. Statistical analyses were performed using Python 3.10.8.

## Data Availability

The datasets generated and/or analyzed during the study, as well as codes and ML models, are not publicly available due to commercial confidentiality and privacy restrictions.

## Author Contributions

Conceptualization: M.M., K.B.

Data curation: M.M., K.K., M.R., K.B.

Formal analysis: M.M., K.K., M.R.

Funding acquisition: K.B.

Investigation: M.U.M., E.B., M.E.G., I.S.S., S.K.

Methodology: M.M., K.B.

Project administration: M.M., K.B.

Resources: M.U.M., E.B., M.E.G., I.S.S., S.K.

Software: K.K., M.R. Supervision: M.M.

Validation: K.K., M.R.

Visualization: M.M.

Writing - original draft: M.M., K.B.

Writing - review & editing: All co-authors.

