## Supplementary S1 for "Real-World Adaptation and External Validation of Machine-Learning Impedance Spectroscopy for Immediate Postpartum Detection of Obstetric Anal Sphincter Injuries"

| Section and topic | No. | STARD-AI item | |
|---|---|---|---|
| Title or abstract | | | |
| | 1† | Identification as a study reporting AI-centered diagnostic accuracy and reporting at least one measure of accuracy within title or abstract | The title notes "Machine-Learning" and the abstract reports the performance metrics such as accuracy. |
| Abstract | | | |
| | 2 | Structured summary of study design, methods, results and conclusions (for specific guidance, see STARD for Abstracts) | The abstract contains all necessary elements, but without explicitly structural headers (because of journal requirements). |
| Introduction | | | |
| | 3† | Scientific and clinical background, including the intended use of the index test, whether it is novel or an established index test and its integration into an existing or new workflow, if applicable | Introduction, positioning the tool as an objective adjunct to bedside clinical examination. |
| | 4 | Study objectives and hypotheses | Stated at the end of the Introduction. |
| Methods | | | |
| Study design | 5 | Whether data collection was planned before the index test and reference standard were performed (prospective study) or after (retrospective study) | Phase 1 and Phase 3 were prospective studies; Phase 2 involved secondary use of routinely collected post-market surveillance data. |
| Ethics | 6* | Formal approval from an ethics committee. If not required, justify why. | Information provided in section 4.1. Study Design and Overview. |
| Participants | 7† | Eligibility criteria: listing separate inclusion and exclusion criteria in the order that they are applied at both participant level and data level | The exclusion and inclusion criteria were prepared for each phase. However, in the manuscript, only the phase 3 criteria were included in section 4.5. |
| | 8 | On what basis potentially eligible participants were identified (such as symptoms, results from previous tests and inclusion in registry) | Identified based on risk factors or diagnostic uncertainty post-vaginal delivery. |
| | 9 | Where and when potentially eligible participants were identified (setting, location and dates) | Depended on phase:<br>Phase 1 in Kosice, Brno, Prague, Warsaw and Leon.<br>Phase 2 in 7 hospitals in Poland, Spain and Austria.<br>Phase 3 in Holy Family Specialist Hospital in Warsaw. |

| | | | |
|---|---|---|---|
| | 10 | Whether participants formed a consecutive, random or convenience series | Stated as prospectively and consecutively recruited. |
| Dataset | 11* | Source of the data and whether they have been routinely collected, specifically collected for the purpose of the study or acquired from an open-source repository | Phase 1/3 as prospective studies and Phase 2 as post-market surveillance data. |
| | 12* | Who undertook the annotations for the dataset (including experience levels and background) and how (within the same clinical context or in a post hoc fashion), if applicable | In Phase 3 reference standard was assessed by a single expert ultrasonographer with >25 years of experience using Sultan criteria. |
| | 13* | Devices (manufacturer and model) that were used to capture data; software (with version number) used to engineer the index test, highlighting the intended use | ONIRY System |
| | 14* | Data acquisition protocols (for example, contrast protocol or reconstruction method for medical images) and details of data preprocessing, in sufficient detail to allow replication | A low-amplitude sinusoidal alternating current (<1 mA) is applied across a frequency range of 1–100 kHz, generating measurements of impedance magnitude, resistance, reactance and phase shift. |
| Test methods | 15a | Index test, in sufficient detail to allow replication | Hardware is detailed, but specific MLP architecture/ hyperparameters are intentionally withheld due to commercial confidentiality. |
| | 15b* | How the index test was developed, including any training, validation, testing and external evaluation, detailing sample sizes, when applicable | Described in sections 4.3, 4.4 and 4.5 |
| | 15c | Definition of and rationale for test positivity cutoffs or result categories of the index test, distinguishing prespecified from exploratory | Models decision threshold was set during development to optimize balanced accuracy, while maintaining the prespecified target sensitivity of >80%. In the phase 3 this threshold was not changed.<br>Final positive test result was obtain if device retuned REFER output. |
| | 15d* | The specified end-user of the index test and the level of expertise required of users | Examinations were performed by trained midwives and physicians. |
| | 16a | Reference standard, in sufficient detail to allow replication | Three-dimensional EAUS. |

| | | | |
|---|---|---|---|
| | 16b | Rationale for choosing the reference standard (if alternatives exist) | Evaluated according to Sultan criteria. |
| | 16c | Definition of and rationale for test positivity cutoffs or result categories of the reference standard, distinguishing prespecified from exploratory | Evaluated according to Sultan criteria. |
| | 17a | Whether clinical information and reference standard results were available to the performers or readers of the index test | Only clinical information |
| | 17b | Whether clinical information and index test results were available to the assessors of the reference standard | No, except for Phase 3: the person performing EAUS was not blinded to prior clinical and ONIRY results. |
| Analysis | 18 | Methods for estimating or comparing measures of diagnostic accuracy | Described in section 4.6. Statistical Analysis. |
| | 19 | How indeterminate index test or reference standard results were handled | Data were checked during data processing and quality control to ensure they met the typical range. |
| | 20 | How missing data on the index test and reference standard were handled | Participants without EAUS follow-up were properly excluded from the paired analysis. |
| | 21 | Any analyses of variability in diagnostic accuracy, distinguishing prespecified from exploratory | Provided in “Results” section. |
| | 22 | Intended sample size and how it was determined | Based on patients availability |
| | 23* | Details of any performance error analysis and algorithmic bias and fairness assessments, if undertaken | Phase 2 of the study |
| Results | | | |
| Participants and dataset | 24 | Flow of participants, using a diagram | Not applied |
| | 25† | Baseline demographic, clinical and technical characteristics of training, validation and test sets, if applicable | Provided in “Results” section. |
| | 26a | Distribution of severity of disease in those with the target condition | Provided in “Results” section. |
| | 26b | Distribution of alternative diagnoses in those without the target condition | Not applicable |
| | 27 | Time interval and any clinical interventions between index test and reference standard | Provided in sections 4.3, 4.4 and 4.5. |
| | 28* | Whether the datasets represent the distribution of the target condition that one would expect from the intended use population | No, the datasets enriched the minority class. |
| | 29* | For external evaluation on an independent dataset, an assessment of how this | Described in results |

| | | | |
|---|---|---|---|
| | | differs from the training, validation and test sets | |
| Test results | 30 | Cross-tabulation of the index test results (or their distribution) by the results of the reference standard | Provided as Table 1. |
| | 31 | Estimates of diagnostic accuracy and their precision (such as 95% confidence intervals) | Provided in Table 1. and results section |
| | 32 | Any adverse events from performing the index test or the reference standard | Adverse events were not reported |
| Discussion | | | |
| | 33 | Study limitations, including sources of potential bias, statistical uncertainty and generalizability | Provided in Discussion section (small sample, single center, loss to follow-up, unblinded evaluator). |
| | 34 | Implications for practice, including the intended use and clinical role of the index test | Positioned as a supplementary tool, not a replacement for clinical examination |
| | 35* | Ethical considerations and adherence to ethical standards associated with the use of the index test and issues of fairness | Addressed via ethics committee approvals and exploratory descriptive performance by subgroups. |
| Other information | | | |
| | 36 | Registration number and name of registry | Phase 1 is registered (NCT04903977). Phase 3, as a non-interventional observational study, was not registered. |
| | 37 | Where the full study protocol can be accessed | Phase 1: the study is described in the ClinicalTrials.gov record (NCT04903977); the full study protocol is not publicly available. Phase 3: the study protocol is not publicly available, as stated in the corresponding publication [13]. |
| | 38 | Sources of funding and other support; role of funders | Addressed in the Funding section. |
| | 39* | Commercial interests, if applicable | Addressed in the Competing Interests section. |
| | 40a* | Availability of datasets and code, detailing any restrictions on their reuse and repurposing | Unavailable due to commercial confidentiality. |
| | 40b* | Whether outputs are stored, auditable and available for evaluation, if necessary | The data are stored in case of audit. |
